# Parental experience of having a child with hypoxic ischaemic encephalopathy: a qualitative study

**DOI:** 10.1101/2025.07.11.25331288

**Authors:** Alexandra Bache, Alastair Sutcliffe, Monica E. Lemmon, Carrie Williams, Chris Gale, Sarah Land, Philippa Rees

## Abstract

**Objective:** To explore hypoxic ischaemic encephalopathy (HIE) families’ experiences of care in the NHS and the impact of HIE on families.

**Design:** Geographically maximum variation sampled semi-structured interviews (n=28) were conducted with parents of infants (born 2010-2024) who underwent therapeutic hypothermia for HIE. Data were analysed with reflexive thematic analysis.

**Setting:** Parents were recruited from across the United Kingdom (UK), covering 84% (11/12) of the UK’s regional neonatal networks, known as Operational Delivery Networks.

**Findings:** Three themes with eight sub-themes were generated from the interview data. 1) The life-changing diagnosis of HIE: Parents described loss of stability & opportunity to parent, ongoing mental turmoil, and how the diagnosis led to transformation. 2) Balancing hope with facts: Parents unpacked how treasured their child is, the tension between hope and loss they experienced and feelings of being kept in the dark. 3) Struggling to meet their child’s needs: Parents outlined deficiencies in care infrastructure, and battling disability-based discrimination.

**Conclusions:** This study highlights the profound and life-changing impact of HIE on families. Parents described cherishing their children and experiencing personal growth. However, many also characterised how challenges were intensified by disability-based discrimination, poor communication and gaps in support across health, education, and social care systems.

To prevent further trauma and support family wellbeing, this work identifies priority improvement areas. Embedding trauma-informed care, strengthening transparent and sensitive communication around prognostic uncertainty, and improving care co-ordination will help families feel seen, heard, and supported throughout their journey.

**SUMMARY:** *What is already known on this topic:*

- Single centre studies demonstrate receiving a HIE diagnosis and having a baby undergo therapeutic hypothermia treatment is traumatic for families.
- Little is known about the experiences of families affected by HIE in the NHS, particularly after neonatal discharge.

*What this study adds:*

- This UK-wide study demonstrates the profound and life changing impact of HIE on families.
- Parents cherish their children and may experience personal growth after HIE; however, family well-being can be undermined by disability-based discrimination, poor communication and inadequately resourced health, education and social care.

*How this study might affect research, practice or policy:*

- These findings support embedding trauma-informed care, encouraging transparent yet sensitive communication around diagnosis and prognosis, and improving care co-ordination to help families.

## INTRODUCTION

Hypoxic ischaemic encephalopathy (HIE) is the leading cause of brain injury amongst term infants, with moderate-severe HIE affecting 1.6-1.9 per 1000 live births annually in England^1^. Since 2010, the National Institute for Health and Care Excellence guidelines have recommended therapeutic hypothermia for moderate-severe HIE as it prevents one case of death or disability for every seven babies treated^2^.

The care pathway for infants with HIE is traumatic for parents. Due to the emergency nature of hypoxia, parents suddenly face uncertainty about their child’s life-expectancy and neurological outcome^3^ ^4^. As care is organised into regional neonatal Operational Delivery Networks (ODNs), babies born outside tertiary centres will require rapid transfer to a different hospital to initiate therapeutic hypothermia. Most families are unfamiliar with HIE and the umbrella term neonatal encephalopathy^5^. Families may encounter further emotional, psychological and financial demands raising children with disabilities and specialised care needs ^6^.

Existing studies in the US and Sweden have largely focused on parental experiences of therapeutic hypothermia^4^ ^7–9^ and prognostic communication^10^ ^11^. These highlight the range of emotions parents experience^3^ ^7^ including chaos and uncertainty^11^ ^12^, a loss of normalcy^13^ and also ongoing parent psychological effects^14^. However, little is known about how families experience HIE care during and after their baby’s time in the neonatal unit, or how this diagnosis affects their lives moving forward^14^ ^15^.

## Methods

We conducted a qualitative study to explore how having a child with HIE impacts families and families’ experiences of care in the NHS.

### Theoretical framework

To explore lived experiences, we chose an experiential orientation for this qualitative thematic analysis^16^. A critical realist approach underpinned this, which conceptualises that whilst there is a singular independent reality, people perceive this differently^16^ ^17^, influenced by sociological and cultural contexts^16^ ^17^, including the NHS in the UK. We chose the ecological life course approach^18^ as the theoretical framework guiding data collection and analysis to acknowledge environmental and interpersonal impacts.

### Recruitment & sampling

Parents aged 18 or above, who had a UK born child treated for HIE with therapeutic hypothermia as a newborn were eligible. Participants were recruited through a self-selected response to online adverts (Supplement 1) that were shared by Peeps, Hope for HIE, the HIE network, and the CHERUB study. Prospective participants provided data in an expression of interest form (Supplement 2), which formed a sampling frame. Purposive maximum variation sampling was used to maximise diversity amongst participants^19^, across regional ODN, infant and parental characteristics.

### PPI

A workshop held with HIE families as part of the CHERuB study^20^ highlighted the need for this study. It identified families varied experiences including vague HIE communication and insufficient signposting of support.

### Data collection & handling

Interviews were conducted individually online by one of the authors (AB) through Microsoft Teams, following a semi-structured interview guide (Supplement 3) which covered HIE’s impact, neonatal communication and support. Virtual interviews were chosen to help parents to participate alongside caring commitments and to maximise geographic spread^21^. We developed the interview guide with neonatal doctors, families, and HIE charity representatives and piloted it with a qualitative researcher. An iterative approach was used to adjust the guide for clarity and explore emerging areas of interest^22^, including from a parallel survey of families.

Interviews were audio-video recorded and automatically transcribed by Microsoft Teams. Each transcript was checked against the recording for accuracy. Concurrent field notes, reflexive audio and typed notes were documented. Interviews were held between 24^th^ April and 20^th^ June 2024, at the participant’s convenience including evenings and weekends.

Verbatim transcripts and recordings were copied to UCL’s data safe haven, then pseudo-anonymised, with names, places and job roles de-identified with bracketed text.

### Data analysis

AB analysed transcripts using reflexive thematic analysis^16^ ^23^. This included labelling units of meaning or analytic ideas (“coding”). Due to the experiential focus, coding was primarily inductive, data-driven, and semantic, focusing on explicit meaning^16^.

NVivo software was used to organise the data, codes and reflexive memos. Themes were produced by grouping codes to form patterns of shared meaning with a central organising concept^16^, with names and definitions updated progressively. To minimise distress, study finding member checking was not utilised^16^ ^22^.

### Reflexivity

Interviews were conducted by AB, a female research and medical student, characteristics disclosed to participants alongside the research aims. AB had post-graduate qualitative training and no prior relationship with participants.

### Ethical review, consent & safeguarding

Chosen participants were sent study information (Supplement 4) and required to provide signed e-consent (Supplement 5) by the REDCap survey platform to participate. They received a £20 voucher afterwards. During recruitment and interviews, participants were signposted to a support sheet (Supplement 6) reviewed by a Child Bereavement Specialist and a distress protocol (Supplement 7) was followed. UCL Ethics Committee approved the study (project ID: 26861/001).

## RESULTS

### Participants

The interview expression of interest form was completed by 63 participants including one duplication. Thirty participants were interviewed, with two suspected imposter participants excluded. A PID number replaces participants’ names.

Table 1 demonstrates the 28 interview participants’ characteristics, whose children were born between 2010 and 2024. Most participants primarily identified as a mother (n=27, 96.4%), had multiple children (n=18, 64.2%), and with developmental delays following HIE (n=21, 75%). Recorded interviews, excluding the introduction and debrief, lasted on average 59.5 minutes (range 46 -81 minutes). Rather than data saturation, we chose a target sample size of 20-30 interviews^24^.

**Table 1.** Interview participant characteristics, N=28. Note: ethnicity categories have been collapsed to the harmonised high level category standards for reporting.

|  |  | Count (%) | Layer N % |
| --- | --- | --- | --- |
| Relationship with child | Mother | 27 | 96.4% |
|  | Father | 1 | 3.6% |
|  | Other caregiver | 0 | 0.0% |
| Child transferred | Yes | 20 | 71.4% |
|  | No | 8 | 28.6% |
| Child's birth year | 2010-2014 | 5 | 17.9% |
|  | 2015-2019 | 9 | 32.1% |
|  | 2020-2024 | 14 | 50.0% |
| Operational Delivery Network | East Midlands | 3 | 10.7% |
|  | East of England | 2 | 7.1% |
|  | North West | 2 | 7.1% |
|  | London | 3 | 10.7% |
|  | Northern | 0 | 0.0% |
|  | Northern Ireland | 0 | 0.0% |
|  | Scotland | 3 | 10.7% |
|  | Kent, Surrey & Sussex | 2 | 7.1% |
|  | South West | 2 | 7.1% |
|  | Thames Valley & Wessex | 3 | 10.7% |
|  | Wales | 1 | 3.6% |
|  | West Midlands | 5 | 17.9% |
|  | Yorkshire & Humber | 2 | 7.1% |
|  | Not disclosed | 0 | 0.0% |
| Child's outcome of HIE | No developmental concerns or delays | 4 | 14.3% |
|  | Developmental delays/issues | 21 | 75.0% |
|  | Death from HIE | 0 | 0.0% |
|  | Other | 3 | 10.7% |
|  | Prefer not to say | 0 | 0.0% |
| Participant's highest level of education | Secondary school | 0 | 0.0% |
|  | College or A-levels | 4 | 14.3% |
|  | University education | 23 | 82.1% |
|  | Other | 1 | 3.6% |
|  | Prefer not to say | 0 | 0.0% |
| Participant's Age | 18 - 24 | 0 | 0.0% |
|  | 25 - 34 | 9 | 32.1% |
|  | 35 – 44 | 15 | 53.6% |
|  | Over 44 | 2 | 7.1% |
|  | Prefer not to say | 0 | 0.0% |
|  | Not given | 2 | 7.1% |
| Participant's number of children | 1 | 10 | 35.7% |
|  | 2 | 12 | 42.9% |
|  | 3 or more | 6 | 21.4% |
|  | Prefer not to say | 0 | 0.0% |
| Participant's ethnicity | White | 26 | 92.9% |
|  | Mixed/ Multiple ethnic groups | 1 | 3.6% |
|  | Asian/ Asian British | 1 | 3.6% |
|  | Black / African / Caribbean / Black British | 0 | 0.0% |
|  | Other ethnic group | 0 | 0.0% |

### Themes

Eight sub-themes were produced and were grouped under the themes 1) Life changing diagnosis of HIE (Table 2) 2) Balancing hope with facts (Table 3) and 3) Struggling to meet child’s needs (Table 4).

**Table 2.** Theme 1 - Life changing diagnosis of HIE.

| Sub-themes | Supporting interview quotes |
| --- | --- |
| Loss of stability & opportunity to parent | 'Obviously there was a lot of shock and not knowing what was happening and that that kind of really destabilised us'. PID18 |
|  | 'Relationships are not the same. I'm actually in the middle of a divorce.' PID10 |
|  | 'When they took me to go and see him, I didn't know if he was mine.' PID23 |
|  | 'Neonatal unit [...] it's like NASA.' PID02 |
|  | 'The Letby case was within the news [...] we were adamant we would not leave [...] you can't just leave a child in the care of people you don't trust.' PID04 |
|  | 'I heard somebody describe it once, like. [...] an emergency landing [...] all of a sudden you're having to learn to live a life that is completely different to what you thought it would be' PID01 |
|  | 'It meant she wasn't my baby because the nurses would tell you off if you open the incubator too long [...] it's humiliating the way they do it, like you're a naughty school child [...]. Yeah it's not having that control, and that separation' PID03 |
| Ongoing mental turmoil | 'I had this recurring dream of this feeling of being on a table being cut and hurt but not being able to tell anyone.' PID09 |
|  | 'Everybody kept telling you to enjoy the time with your baby [...] while you're just kind of on hold just waiting [...]you had the MRI scan that shows damage [...] But then they tell you that actually it's not a guarantee [...]. And so you're just, kind of up in the air, just kind of waiting for something to happen.' PID06 |
|  | 'People, would you know, say [...] 'Oh well he looks perfectly fine [...] It doesn't look like there's anything wrong with him' [...]. It's like belittling what we went through and what we were still going through [...] but also it's like saying that if he did have cerebral palsy [...] there would be something wrong with him. That really annoys me and upsets me.' PID08 |
|  | 'I'm constantly checking the monitor to see if she's OK at night.' PID19 |
|  | 'Somebody needs to ask them if they're okay? Because I had to, you know, get to a point of being really quite unwell before, you know, anybody sort of asked. [...], perhaps, you know, five years on [...] I wouldn't hold quite so much guilt and sadness about it if somebody had approached the question, you know, then to start the healing.' PID08 |
|  | 'Maybe like a check in at the year, [...] I feel like the first birthday is when [...] things might slow down for parents and they might be able to look back at what happened [...] I think you get so caught up in your child's needs [...] you kind of neglect yourself.' PID12 |
| Parental transformation | 'This HIE event happened. It completely broke me apart [...] I was just a shell [...] I've slowly put myself back together. And, it's just different now, but when I sit and think about it, I prefer this version.' PID10 |
|  | 'A bit more a holistic approach and a holistic view as how people are affected by certain things [...] I get upset about different things [...] I'm quite strong minded about certain things that I probably didn't think about before like accessibility and the way people are spoken about or spoken to.' PID11 |
|  | 'I think it really has completely changed me [...] I had a career before. I kind of left work. I'm down as, I'm a carer [...] So it's made me change my life. My life is now her.' PID12 |
|  | 'I've been much more available to him [her child]. I've had lots more time with him [...] so I've been able to enjoy being a mum. [...] I definitely had my priorities wrong before [...] Everybody's on this treadmill of you must get your child to be the top of the class and winning the football trophies [...] Those things are just not that important.' PID09 |
|  | 'I've shared sort of my struggles with people in a way that I never would have before [...] I've definitely learnt to adapt to that kind of more healthy way of viewing mental struggles [...] I'm still struggling [...] but it's in a much more balanced way [...] I now have ways to cope.' PID13 |

**Table 3.** Theme 2 - Balancing Hope with Facts.

| Sub-themes | Supporting interview quotes |
| --- | --- |
| Treasured child | 'He is exactly the person he needs to be.' PID08 |
|  | 'It is quite demanding, having a complex needs child, and I won't change him for the world like I love my son. He is who he is. He's a cheeky little boy!' PID22 |
|  | 'We're finally enjoying our baby [...] that's such a special thing that it took us so long to get to. [...] try and get to that point as soon as you can.' PID13 |
|  | 'He is worth fighting for.' PID15 |
| Tension of hope and loss | 'I understand the need for the cold hard facts, but [...] there's always room for, for hope and possibilities. [...] After 24 hours was when [...] we started planning his funeral. Because we were given absolutely zero hope whatsoever'. PID16 |
|  | 'I'm probably more of a pessimist [...] but at that point, all you want to do is try and cling to a bit of hope because it's like the darkest day of your life, isn't it, but, yeah I felt, it just all felt very, very factual, clinical.' PID07 |
|  | 'I mourned for the life that I thought I might lose.' PID07 |
|  | 'As much as it's a birthday, it's also an anniversary of something really traumatic that's happened.' PID12 |
|  | 'Mother instincts, that something isn't quite right, [...] that brings you back round to, even like the birth. Like you, you try and say 'Oh I don't think something's right'. 'Oh, no, you're fine, you know.' But it all went wrong. So you kind of want to go with your gut.' PID05 |
|  | 'I was crying and she [a NICU staff member] came over and said, 'Oh no, you need to be the strong one for your wife''. PID |
| Being kept in the dark | 'My son was born. They didn't tell us he had HIE. They didn't explain what it was. They didn't say it could have lifelong implications of severe disability. I think emotionally they kept me in the dark a lot and it's affected my trust of any healthcare professional now.' PID17 |
|  | 'It just felt things were quite vague, you know, around what happened [...] what the tests were showing on her, you know the MRI and things like that [...] it would have been nice to have a bit more information.' PID01 |
|  | 'This brain injury that my son suffered, with this very long, strange name. I've never heard of it. We must be the only family this [has] happened to [...] This is why they don't know what's happening [...] why they think he's going to die because it's so, so incredibly rare.' PID15 |
|  | 'All the doctors, since anytime we've had to go to hospital and mention like medical history and that, none of them know. I had one doctor that knew what HIE was!' PID10 |
|  | 'You know, I've been into the hospital appointment before and someone had 'What is HIE' googled on their screen ((laughing)). That doesn't give you a lot of comfort really.' PID20 |
|  | 'Nobody actually told us that she'd had this HIE event. And it was only from reading it in the paperwork that I was then like, 'oh!'" PID21 |
|  | 'I mean, we tried to get a debrief soon after we were home, but they wouldn't until he'd had his H-S-I-B, HSIB investigation, which takes up to six months.' PID05 |

**Table 4.** Theme 3 - Struggling to meet child’s needs.

| Sub-themes | Supporting interview quotes |
| --- | --- |
| Insufficient care infrastructure | 'I actually sent a complaint e-mail to PALS [patient advice and liaison service] and just said, what does it take [...] to get a piece of supportive equipment that's desperately needed, that's for safety?' PID14 |
|  | 'I lost the feeling in both my legs, which was from holding [child's name] for literally 20 hours a day [...] because it was the only way she would tolerate feed and not choke [...] So that was when social said they might try and help us. Unfortunately, then my legs improved, so no help.' PID04 |
|  | 'I'm terrified of taking him to appointments [...]. The amount of times I've made like emergency stops in places I shouldn't to literally run around the side of the car to make sure he's not choking, is very difficult.' PID22 |
|  | 'Trying to get his needs met: so his physical needs, therapy needs, social needs, it's everything is a constant battle [...] He has a specialist bed, a specialist indoor chair, a specialist buggy, a specialist bath seat. Splint for his legs. He's just got his EHCP [education and health care plan]. He needed a one-to-one for nursery, he's gonna need a one-to-one for school speech and language therapy. Physiotherapy, basically everything. [...] Everyone's so stretched and underfunded and understaffed. That you have to really fight tooth and nail to have his needs met [...] for what he's legally entitled to.' PID16 |
| Battling disability-based discrimination | 'Like from the minute that [child's name] was born, we were very much told all the things he may not do. And very few people told us all the things that he would do [...] like almost like barriers put up...' PID08 |
|  | 'They were very negative about disability. Broadly you know "I'm so sorry for your loss. Your child might have a disability". You know, it's like the worst thing that could ever happen to you is your child being disabled. And no, looking back [...], it's not the worst thing that ever happens to you.' PID02 |
|  | 'Because he has a significant brain injury [...] he has been written off by a few professionals, especially when it comes to the epilepsy side of things [...] that really distresses us as parents that. He is worth fighting for.' PID15 |
|  | 'I don't think they want her in there [...] she needs a standing frame [...] the nursery is huge. They've got plenty of room for it and they're refusing. So that's a battle.' PID14 |
|  | 'The NHS don't have time for children like [child's name] at all, so he's completely excluded from society in every way possible.' PID02 |
|  | 'It's not just I have to deal with institutional racism, the unconscious bias [...] there's so many layers [...] when you're in pain, it's not being taken seriously [...] when your child needs care oh you're being aggressive.' PID03 |
|  | 'And then socially we can't go the park - we can go the park but she can't go on any of the swings.' PID14 |
|  | 'It just makes you more aware when you go somewhere, and you think, do you know what would be really great here, a Changing Places toilet? [...] Everywhere you go should be, you know, in this day and age, accessible for everybody. But unfortunately it's not.' PID24 |

#### Life-changing diagnosis of HIE (Table 2)

##### Loss of stability & opportunity to parent

Loss of stability describes the far-reaching consequences of the HIE event, in parents’ plans and relationships. One parent likened this to ‘*an emergency landing’* (PID01). For some, friends and family gave stability, whilst others found friendships dissolved and the relationship with their partner broke down.

Initial losses of opportunity to parent included delayed milestones of first meeting, feeding and holding their baby. Parents felt under scrutiny in the unfamiliar neonatal unit ‘*like NASA’* (PID02), and were admonished, for example when opening the incubator too long: *‘It’s humiliating the way they do it, like you’re a naughty school child’ (*PID03), contributing to the loss of control. One parent highlighted the Lucy Letby media coverage’s^25^ impact: ‘*we would not leave […] you can’t just leave a child in the care of people you don’t trust’* (PID04).

##### Ongoing mental turmoil

The traumatic nature of the initial HIE event took a protracted mental toll on parents. Future uncertainty continued into their child’s adolescence. *“You’ve gone through a very big traumatic experience to ‘Ok you can go home, now’ and you go home and you’re just waiting’* (PID05). Parents felt *‘on hold’* (PID06), *‘almost like a grenade’s been thrown’* (PID07) as they awaited the development of possible neurological symptoms. Comments their child ‘*look[ed] perfectly fine’* were difficult for some parents to hear. Parents felt this was *‘belittling what [they] went through […] but also it’s like saying that if [they] did have cerebral palsy […] there would be something wrong with [their child]’* (PID08). Some parents extensively researched HIE but found this fuelled worry. Many parents were diagnosed with post-traumatic stress disorder (PTSD) and anxiety, experiencing guilt and hypervigilance. Some experienced nightmares, a *‘recurring dream of this feeling of being on a table being cut but not being able to tell anyone’* (PID09). Parents felt earlier support might have helped, reflecting *‘perhaps, you know, five years on […] I wouldn’t hold so much guilt and sadness about it’* (PID08).

##### Parental transformation

Some felt the HIE event triggered positive personal change. One parent described becoming *‘an entirely different person’*, having been ‘*broke apart’* by the HIE event, she says I *‘slowly put myself back together. And it’s just different now, but when I sit and think about it, I prefer this version’* (PID10). Another developed a more ‘*holistic approach’* at work (PID11). Some parents needed to redefine their identity. *‘I had a career before. [..] I’m a carer […] My life is now her’* (PID12). Some mothers felt the injustice of being expected to sacrifice their career. However, others saw the benefits in being *‘much more available […] so I’ve been able to enjoy being a mum’* (PID09), with priorities shifted from their child’s external success to happiness. Parents developed emotional coping skills with a *‘more healthy way of viewing mental struggles’* (PID13).

#### Balancing hope with facts (Table 3)

##### Treasured child

Parents described their love of spending time with their child, of whom they were proud and accepting. *‘He is exactly the person he needs to be’ (PID08).* Within this they acknowledged the difficulty of their situation*. ‘It is quite demanding, having a complex needs child, and I won’t change him for the world like I love my son. He is who he is. He’s a cheeky little boy’* (PID22). Parents appreciated NICU staff who helped them interact, through bathing or stroking their baby. As advocates, parents emphasised their child being *‘worth fighting for’* (PID15).

#### Tension between hope and loss

During the neonatal unit admission, parents felt being given hope was essential amongst the ‘*cold hard’* clinical facts but acknowledged the difficulties doctors’ face in offering hope amidst uncertainty. *“After 24 hours […] we started planning his funeral. Because we were given absolutely zero hope”* (PID16). Many parents were grieving, regardless of their child’s outcome, as one parent says, *‘I mourned for the life that I thought I might lose’* (PID07). Their child’s birthday was *‘an anniversary of something really traumatic’* (PID12). In some incidents professionals negatively challenged parents’ emotions. This sometimes invoked memories that their concerns were not acted on during birth when they were reassured *‘“You’re fine you know”. But it all went wrong’*(PID05). One crying parent was told ‘*you need to be strong one for your wife’* (PID).

##### Being kept in the dark

*S*ome parents felt information about their child was withheld and only emerged through clinical investigations. This lack of transparency damaged trust in the NHS. *‘They didn’t tell us he had HIE. […] emotionally they kept me in the dark a lot and it’s affected my trust of any healthcare professional’ (*PID17). Additionally, parents found HIE information ‘*quite vague’* (PID01), sometimes only learning this diagnosis through discharge paperwork. Parents then found HIE was unfamiliar to health visitors, midwives, general practitioners and emergency doctors: ‘*I had one doctor that knew what HIE was!*’ (PID10). Further a few parents found placental analysis results were not readily shared, which extended feelings of guilt they associated with the birth injury and removed choice in deciding whether to have more children.

#### Struggling to meet child’s needs (Table 4)

##### Insufficient care infrastructure

Many children had additional health, educational and social needs. However, the lack of supportive infrastructure frustrated parents, forcing them into a costly, time-consuming litigation process. One parent asked PALS [patient advice liaison service] *‘what does it take […] to get a piece of supportive equipment that’s desperately needed? That’s for safety’* (PID14). Parents were providing full time care, including holding their child for *‘literally 20 hours a day [..] because that was the only way she would tolerate feed and not choke’* (PID04) but received no further support. Several parents described the stress of making emergency stops whilst travelling to appointments with their child at risk of choking, due to lack of appropriate seating. Necessary equipment included specialist beds, chairs, buggies, bath seats, and leg splints but parents felt forced to ‘*fight tooth and nail’ (PID16)* to access these.

##### Battling disability-based discrimination

Parents felt they encountered discrimination against their child in educational and healthcare settings, which failed to make accessibility adaptations. *‘I don’t think they want her in [nursery] […] she needs a standing frame […]. They’ve got plenty of room for it and they’re refusing’ (*PID14). Several children had left school due to a lack of support. Experiences of disability-based discrimination combined with others forms of discrimination like racism: ‘*It’s not just I have to deal with institutional racism, the unconscious bias […] there’s so many layers’* explaining *‘when you’re in pain, it’s not being taken seriously […] when your child needs care oh you’re being aggressive’(PID03)*.

Prognostic information was sometimes felt to be negative and framed by a negative view of disability, ‘*we were very much told all the things he may not do. And very few people told us all the things that he would do*’ (PID08). Parents were hurt by the sense their child was written off by the NHS, with financial savings prioritised over their child’s care. ‘*The NHS don’t have time for children like [child’s name] at all, so he’s completely excluded from society’* (PID02). For some families, insufficient care infrastructure was perceived as systemic discrimination against their child with a disability.

## DISCUSSION

Our study findings demonstrate the profound and life-changing impact of HIE on families. Parents cherish their children and may experience personal growth. However, parents found disability-based discrimination, guarded and insensitive communication and inadequately resourced health, education and social resources made accessing essential care challenging.

### Strengths & Limitations

A major strength is this study’s geographic representation across the UK, with participants from 84% (11/12) of regional neonatal networks. Independence of researchers from the clinical team may have reduced the power asymmetry^26^ enabling disclosure of negative or discriminatory experiences. Although the primary researcher’s lack of lived parenthood experience could mean nuance was missed^27^, it may have facilitated parents’ openness, as parents often positioned their experiences relative to other parents’. Perceiving the researcher as an ‘outsider’, may have increased parents’ confidence in their own experience^27^. Furthermore, this study’s larger sample size^15^ ^13^ provides strength, through highlighting shared and divergent experiences across families in different circumstances, enabled by purposive maximum variation sampling^19^.

A study limitation is the recruitment’s reliance on support groups. While successfully reaching many parents, it may have introduced selection bias. Notably, fathers and bereaved parents were underrepresented despite targeted recruitment. Additionally, White British (n = 24, 85.7%) and university educated (n = 23, 82.1%) participants were overrepresented.

While recall bias may be considered a limitation, parents’ subjective perception and memory of events is critically important, as it constitutes their lived experience which influences support needs. Furthermore, the £20 voucher provided may have introduced motivation bias, resulting in the attempted participation of imposter respondents who were excluded.

### Context with literature

The theme of “loss of stability” reported in this study intersects with the concept of chaos previously described^3^. Additionally, previously highlighted "repeated losses" or "longitudinal grief"^12^ themes, highlight grief around HIE as an ongoing process. Further loss of opportunity to parent has been previously identified^28^.

The theme “being kept in the dark” aligns with existing data suggesting families experience unclear neurologic prognostication^29^ and fragmented communication^10^. Mistrust may develop when nonspecialist caregivers provide initial neonatal information^10^. Amongst the theoretical discussions around HIE and neonatal encephalopathy terminology^5^, this study emphasises the real-life need to explain diagnoses to families in a timely manner to maintain their trust and enable them to access support^5^. This study adds that withholding placental analysis results, can augment harm by limiting parents’ opportunities to make informed reproductive choices.

Furthermore, while aspects of the "ongoing mental turmoil" theme—like the burden of uncertainty—have been reported elsewhere^10^ ^13^, this study uniquely emphasises that negatively framing disability contributes to healthcare and education exclusion alongside affecting families directly. VanPuymbrouck, Friedman, and Feldner^30^ note many healthcare professionals harbour implicit biases against individuals with disabilities^30^, often stemming from a lack of knowledge and a tendency to dehumanise them^31^. This shows the far-reaching impact of negative narratives on families affected by HIE.

The “parental transformation” theme builds on a previously identified theme of positive adaptation^15^, but further adds that parents’ priorities may be altered and identity reformed. This aligns with the findings from a systematic review of the positive aspects of parenting a child with intellectual difficulties^6^ which also identifies the child as a source of happiness and fulfilment, overlapping with the “treasured child” theme.

#### Research implications

Future research should explore the experiences of underrepresented HIE populations. This includes people from minority ethnic groups, bereaved families as well as fathers, siblings and grandparents. To better counsel families, methods and training to communicate uncertainty and neurological outcomes, including the role of prognostic indicators, warrant further investigation.

### Policy & practice implications

The importance of perinatal brain injuries, like HIE, is recognised in current UK policy, including the Department of Health and Social Care’s national maternity ambition^32^. The consequences of these injuries on children, families, and society highlight the urgent need to improve care pathways.

Therefore, eight core HIE care principles grounded in parent experience are proposed. These include: (1) provide stability, (2) deliver trauma-informed care, (3) ensure transparent timely communication of HIE diagnosis, (4) provide training on language used to communicate about disability, (5) simplify access to care, (6) offer psychological support, and (7) resource parents through change. Overarching (8) is the goal of demonstrating care for the child and empowering families to love, bond with, and cherish their child (see Table 5).

**Table 5.** Policy and practice recommendations.

|  | Care principle | Practical recommendations |
| --- | --- | --- |
| Care for child | Provide stability | • Enable parents to stay near their child to minimise parental distress and facilitate trauma recovery<sup>34 39</sup><br>• Adopt other methods of connection like live video<sup>35</sup> where parents are unable to be near infant<br>• Maximise parental involvement in infant care like first bathing, dressing and nappy change as well as medication delivery<sup>39</sup> |
|  | Deliver trauma informed care | • Provide trauma informed care training for neonatal staff, health visitors, GPs, and other professionals<sup>37</sup><br>• Offer choice where possible (for example in infant clothing) and acknowledge parents' experiences nonjudgmentally<br>• Train staff to view and describe disability in a balanced way<sup>30</sup> |
|  | Transparent communication | • Describe HIE as likely diagnosis, whilst acknowledging uncertainty, as soon as possible. Explain clearly terminology such as neonatal encephalopathy and HIE<br>• Emphasise open, transparent communication by offering a discharge meeting as standard care<br>• Provide prompt obstetric debrief offering discussion of risk in potential future pregnancies<br>• Ensure legal investigations do not preclude parents being fully informed of clinical events or results<br>• Increase awareness of HIE amongst healthcare professionals |
|  | Simplify care access | • Consider assessing children to determine if a key worker<sup>40</sup> (care co-ordinator & advocate) would help<br>• Allocate more funding to meet families' care and therapy needs, including funding carers, supportive car seats/accessible vehicles, and home equipment such as specialist bath seats, indoor chairs and buggies<br>• Prioritise accessibility in community buildings and spaces, including playgrounds and facilities like Changing Places toilets |
|  | Offer psychological support | • Link parents to NICU or HIE peer support<sup>38</sup><br>• Signpost parents to relevant charities<br>• Provide uniform psychological screening and support in the NICU<sup>14 39</sup> |
|  | Resource parents in change | • Offer a parental check-in<sup>14</sup>, for example with the GP around a year after the child's birth<br>• Support parents in accessing neonatal care pay and leave, and extending leave further where possible |

Stability and safety are key components of trauma treatment models^33^. These can be increased by enabling parents to stay near their child ^34^ or offering live video connection^35^. Secondly, healthcare settings may re-traumatise parents through removing choice, minimising their experiences, or emotional insensitivity^36^ ^37^.

Therefore, all healthcare professionals may benefit from trauma-informed care training. Recent press coverage of maternal and neonatal care failings may further undermine trust ^25^, emphasising the importance of open communication. A discharge meeting should always be offered, with prompt obstetric debrief to discuss the birth and future pregnancy implications.

Given peer support facilitates parental role attainment,^38^ and parental empowerment is a feature of family integrated care^39^, units should signpost to sources like HIE charities. Improving neonatal and community care transition and increasing inter-specialty learning will benefit families and professionals.

In conclusion, this study highlights priority areas for improving support for families affected by HIE. These findings support embedding trauma-informed care, encouraging transparent and sensitive communication and improving care co-ordination to create a more healing environment.

## Data Availability

The study data including audio-visual recordings will not be publicly available due to containing sensitive and identifiable data. Metadata will be made available.

## ACKNOWLEDGEMENTS

Thank you to Peeps, Hope for HIE and the HIE network for their help with recruitment and feedback.

Thank you to Dr Celine Lewis for her valuable input throughout the study.

Thank you to Rachel Cooke, Great Ormond Street Hospital Bereavement Service Manager, for her help reviewing family resources and providing psychological debrief.

## COMPETING INTERESTS

CG has received salary support from the Medical Research Council and grant funding paid to his institution from the National Institute for Health Research (NIHR), Action Medical Research, the Canadian Institute for Health Research, the Medical Research Council, and Chiesi Pharmaceuticals. CG has also received payment of travel and accommodation to attend educational meetings from Chiesi Pharmaceuticals. SL is the Co-Founder and Charity Manager at Peeps, a charity supporting those affected by HIE.

## FUNDING

A £600 stipend to cover thirty £20 vouchers was provided by the UCL MRes course.

This study is an extension of the CHERuB study, which is funded by the National Institute for Health and Care Research (NIHR301457) and the NIHR GOSH BRC. The views expressed are those of the author(s) and not necessarily those of the NHS, the NIHR or the Department of Health and Social Care.

## Notes

### Funding Statement

A 600 pound stipend to cover thirty 20 pound vouchers was provided by the UCL MRes course.
This study is an extension of the CHERuB study, which is funded by the National Institute for Health and Care Research (NIHR301457) and the NIHR GOSH BRC. The views expressed are those of the author(s) and not necessarily those of the NHS, the NIHR or the Department of Health and Social Care.

### Author Declarations

University College London Research Ethics Committee of University College London gave ethical approval for this work (ID 26861/001).

## REFERENCES

1. Gale C, Statnikov Y, Jawad Sea. Correction: Neonatal brain injuries in England: population-based incidence derived from routinely recorded clinical data held in the National Neonatal Research Database. Archives of Disease in Childhood - Fetal and Neonatal Edition 2021;106(3):e1. doi: 10.1136/archdischild-2017-313707corr1

2. Jacobs SE, Berg M, Hunt R, et al. Cooling for newborns with hypoxic ischaemic encephalopathy. Cochrane Database Syst Rev 2013;2013(1):Cd003311. doi: 10.1002/14651858.CD003311.pub3 [published Online First: 20130131]

3. Heringhaus A, Blom MD, Wigert H. Becoming a parent to a child with birth asphyxia-From a traumatic delivery to living with the experience at home. International Journal of Qualitative Studies on Health and Well-being 2013;8 doi: 10.3402/qhw.v8i0.20539 [published Online First: 20130430]

4. Nassef SK, Blennow M, Jirwe M. Parental viewpoints and experiences of therapeutic hypothermia in a neonatal intensive care unit implemented with Family-Centred Care. Journal of Clinical Nursing 2020;29(21-22):4194–202. doi: 10.1111/jocn.15448 [published Online First: 20200820]

5. Molloy EJ, Branagan A, Hurley T, et al. Neonatal encephalopathy and hypoxic– ischemic encephalopathy: moving from controversy to consensus definitions and subclassification. Pediatric Research 2023;94(6):1860–63. doi: 10.1038/s41390-023-02775-z

6. Beighton C, Wills J. How parents describe the positive aspects of parenting their child who has intellectual disabilities: A systematic review and narrative synthesis. J Appl Res Intellect Disabil 2019;32(5):1255–79. doi: 10.1111/jar.12617 [published Online First: 20190520]

7. Bäcke P, Hjelte B, Hellström Westas L, et al. When all I wanted was to hold my baby-The experiences of parents of infants who received therapeutic hypothermia. Acta Paediatr 2021;110(2):480–86. doi: 10.1111/apa.15431 [published Online First: 20200724]

8. Lemmon ME, Donohue PK, Parkinson C, et al. Parent Experience of Neonatal Encephalopathy: The Need for Family-Centered Outcomes. Journal of Child Neurology 2016;32(3):286–92. doi: 10.1177/0883073816680747

9. Thyagarajan B, Baral V, Gunda R, et al. Parental perceptions of hypothermia treatment for neonatal hypoxic-ischaemic encephalopathy. J Matern Fetal Neonatal Med 2018;31(19):2527–33. doi: 10.1080/14767058.2017.1346074 [published Online First: 20170711]

10. Lemmon ME, Donohue PK, Parkinson C, et al. Communication Challenges in Neonatal Encephalopathy. Pediatrics 2016;138(3) doi: 10.1542/peds.2016-1234 [published Online First: 20160803]

11. Craig AK, Gerwin R, Bainter J, et al. Exploring Parent Experience of Communication About Therapeutic Hypothermia in the Neonatal Intensive Care Unit. Advances in Neonatal Care 2018;18(2):136–43. doi: 10.1097/anc.0000000000000473

12. Craig AK, Gerwin R, Bainter J, et al. Exploring parent expectations of neonatal therapeutic hypothermia. Journal of Perinatology 2018;38(7):857–64. doi: 10.1038/s41372-018-0117-8 [published Online First: 20180508]

13. Nassef SK, Bohlin MB, Jirwe M. Experiences of parents whose school-aged children were treated with therapeutic hypothermia as newborns: A focus group study. Nursing Open 2023;10(11):7411–21. doi: 10.1002/nop2.1994 [published Online First: 20230926]

14. Kokkonen Nassef S, Blennow Bohlin M, Jirwe M. Experiences of parents whose school-aged children were treated with therapeutic hypothermia as newborns: A focus group study. Nursing Open 2023;10(11):7411–21. doi: 10.1002/nop2.1994 [published Online First: 20230926]

15. Kramer N. Parents of Children Who Had Hypoxic-Ischemic Encephalopathy: A Mixed-Methods, Exploratory Study: Duquesne University, 2017.

16. Braun V, Clarke V. Thematic analysis : a practical guide London: SAGE Publications Ltd 2022.

17. Pilgrim D. Some implications of critical realism for mental health research. Social Theory & Health 2014;12(1):1–21. doi: 10.1057/sth.2013.17

18. Tomlinson M, Hunt X, Daelmans B, et al. Optimising child and adolescent health and development through an integrated ecological life course approach. BMJ 2021;372:m4784. doi: 10.1136/bmj.m4784

19. Palinkas LA, Horwitz SM, Green CA, et al. Purposeful Sampling for Qualitative Data Collection and Analysis in Mixed Method Implementation Research. Administration and Policy in Mental Health and Mental Health Services Research 2015;42(5):533–44. doi: 10.1007/s10488-013-0528-y

20. Rees P, Gale C, Battersby C, et al. Childhood Health and Educational outcomes afteR perinatal Brain injury (CHERuB): protocol for a population-matched cohort study. BMJ Open 2024;14(8):e089510. doi: 10.1136/bmjopen-2024-089510

21. Krouwel M, Jolly K, Greenfield S. Comparing Skype (video calling) and in-person qualitative interview modes in a study of people with irritable bowel syndrome – an exploratory comparative analysis. BMC Medical Research Methodology 2019;19(1):219. doi: 10.1186/s12874-019-0867-9

22. Grbich C. Qualitative data analysis: An Introduction 2nd ed. London SAGE Publications 2013.

23. Braun V, Clarke V. Reflecting on reflexive thematic analysis. *Qualitative Research in Sport*, Exercise and Health 2019;11(4):589–97. doi: 10.1080/2159676X.2019.1628806

24. Braun V, Clarke V. To saturate or not to saturate? Questioning data saturation as a useful concept for thematic analysis and sample-size rationales. *Qualitative Research in Sport*, Exercise and Health 2021;13(2):201–16. doi: 10.1080/2159676X.2019.1704846

25. Iacobucci G. Lucy Letby is found guilty of attempting to murder premature baby after retrial. BMJ 2024;386:q1487. doi: 10.1136/bmj.q1487

26. Hunt MR, Chan LS, Mehta A. Transitioning from Clinical to Qualitative Research Interviewing. International Journal of Qualitative Methods 2011;10(3):191–201. doi: 10.1177/160940691101000301

27. Dwyer SC, Buckle JL. The Space Between: On Being an Insider-Outsider in Qualitative Research. International Journal of Qualitative Methods 2009;8(1):54–63. doi: 10.1177/160940690900800105

28. Craig AK, James C, Bainter J, et al. Parental perceptions of neonatal therapeutic hypothermia; emotional and healing experiences. The Journal of Maternal-Fetal and Neonatal Medicine 2020;33(17):2889–96. doi: 10.1080/14767058.2018.1563592 [published Online First: 20190108]

29. Guttmann K, Flibotte J, DeMauro SB, et al. A Mixed Methods Analysis of Parental Perspectives on Diagnosis and Prognosis of Neonatal Intensive Care Unit Graduates With Cerebral Palsy. Journal of child neurology 2020;35(5):336–43. doi: 10.1177/0883073820901412

30. VanPuymbrouck L, Friedman C, Feldner H. Explicit and implicit disability attitudes of healthcare providers. Rehabilitation Psychology 2020;65(2):101–12. doi: 10.1037/rep0000317 [published Online First: 20200227]

31. Ames SG, Delaney RK, Houtrow AJ, et al. Perceived Disability-Based Discrimination in Health Care for Children With Medical Complexity. Pediatrics 2023;152(1):e2022060975. doi: 10.1542/peds.2022-060975

32. Department of Health and Social Care. New NHS programme to reduce brain injury in childbirthlJ[Press release], 2025.

33. Herman JL. Trauma and Recovery. The Psychoanalytic Quarterly 1992;Vol. 64:p. 806–09.

34. Zhang Y, Jiang M, Wang S, et al. Effect of family integrated care on stress in mothers of preterm infants: A multicenter cluster randomized controlled trial. Journal of Affective Disorders 2024;350:304–12. doi: 10.1016/j.jad.2024.01.102

35. Kilcullen ML, Kandasamy Y, Evans M, et al. Parents using live streaming video cameras to view infants in a regional NICU: Impacts upon bonding, anxiety and stress. Journal of Neonatal Nursing 2022;28(1):42–50. doi: 10.1016/j.jnn.2021.03.013

36. Center for Substance Abuse T. SAMHSA/CSAT Treatment Improvement Protocols. Trauma-Informed Care in Behavioral Health Services. Rockville (MD): Substance Abuse and Mental Health Services Administration (US) 2014.

37. Emsley E, Smith J, Martin D, et al. Trauma-informed care in the UK: where are we? A qualitative study of health policies and professional perspectives. BMC Health Services Research 2022;22(1):1164. doi: 10.1186/s12913-022-08461-w

38. Rossman B, Greene MM, Meier PP. The role of peer support in the development of maternal identity for "NICU Moms". Journal of Obstetric, Gynecologic & Neonatal Nursing 2015;44(1):3–16. doi: 10.1111/1552-6909.12527 [published Online First: 20150107]

39. Franck LS, O’Brien K. The evolution of family-centered care: From supporting parent-delivered interventions to a model of family integrated care. Birth Defects Research 2019;111(15):1044–59. doi: 10.1002/bdr2.1521

40. Brenner M, Doyle A, Begley T, et al. Enhancing care of children with complex healthcare needs: an improvement project in a community health organisation in Ireland. BMJ Open Quality 2021;10(1):e001025. doi: 10.1136/bmjoq-2020-001025

