## Supplementary Material for "Parental experience of having a child with hypoxic ischaemic encephalopathy: a qualitative study"

1. Supplement 1 – Advertising material

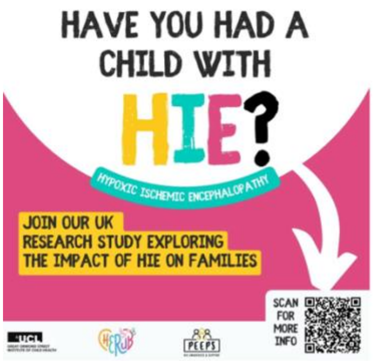

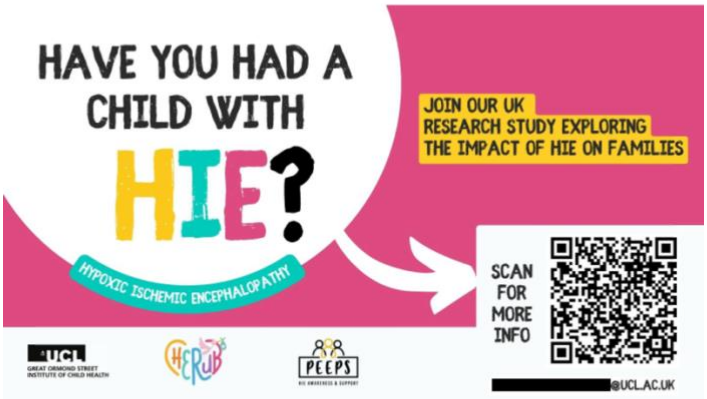

***Supplement Figure 1-1 Recruitment adverts.*** *These are the ethics-approved study adverts which were distributed on social media. Note: the QR codes are no longer functional.*

**1. Email/ Website**

Image: as above

Text:

Have you had a child with HIE?

A UK research study is being done to explore the impact of HIE on families. There are two parts you can choose to be involved in if you are a parent whose child was cooled in the UK

1) An anonymous survey
   access here: [The Impact of HIE on your family Survey]
2) An online interview
   express interest here: [Interview expression of interest]

Taking part is completely your choice. You will be given a £20 thank you voucher for participating in an interview.

Click on the above links to complete the survey and/or express interest. You can also contact the team at for more information.

Note, if the links above do not work, try copying the links below into your web browser:

Survey:
https://redcap.idhs.ucl.ac.uk/surveys/?s=43CDKL9PC4YJMTM3
Express interest in interview:
https://redcap.idhs.ucl.ac.uk/surveys/?s=FWYJX8J4RPFAXXTM

**2. Instagram/ Facebook/ WhatsApp/ LinkedIn**

Image: as above

Text:

Join a UK study exploring the impact of HIE on families.
You can be involved in:
1) an anonymous survey and/or
2) an online interview,
if your child was cooled in the UK.

Taking part is optional.

£20 thank you voucher for interview participation.

Copy this link for the survey:
https://redcap.idhs.ucl.ac.uk/surveys/?s=43CDKL9PC4YJMTM3

Or register interest in an interview:
https://redcap.idhs.ucl.ac.uk/surveys/?s=FWYJX8J4RPFAXXTM

Contact:

**3. X**

Image: as above

Text:

Join a UK study of how HIE impacts families.
Take part in an anonymous survey and/or online interview if your child was cooled in the UK. £20 thank-you voucher for interview.

 for more information

https://redcap.idhs.ucl.ac.uk/surveys/?s=43CDKL9PC4YJMTM3

#### 2. Supplement 2 - Expression of interest

The content of the Interview Expression of Interest is visible here. To see how this appeared to participants see Supplement 8-1A.

### Interview expression of interest

Thank you so much for your interest in taking part in an interview. You are invited to take part if you have had a baby with HIE who was cooled in the UK.

Why is the study been done?

We aim to listen to families who have had a child with HIE in the UK to hear their experiences, such as what the impact has been on their family. This study is part of a Master of Research degree at UCL GOS Institute of Child Health.

This study has been approved by the UCL Research Ethics Committee. Project ID number: 26861/001

What is involved?

We would like to speak to you and listen to your story ("interview" you). This will be online, on Microsoft Teams, at a time that works for you, and will last around one hour.

You will be given a £20 voucher as a thank you for taking part, if you are chosen to be interviewed. A trusted individual may join you for the individual interview, but we are particularly keen to hear your views.

How is this different from the survey?

The survey is a separate part of this research study. You may choose to take part in the interview, or the survey or both if you wish. The survey can be accessed by clicking here.

Note: some of the background and demographic questions are repeated. We understand this is inconvenient to complete twice, however we were keen to maintain the anonymity of the survey and therefore the two responses are separate.

Can I choose to take part?

Yes, taking part is completely your choice. You do not have to take part. Please note, to ensure we listen to a variety of families, it may not be possible to interview all individuals who are interested.

We will select individuals to ensure we listen to families with a varied range of experiences and characteristics.

You can access information about support available here:

Support available for you.pdf

Once you have completed this form we will be in touch with more information. If you would prefer to contact us in advance you can. Contact Alexandra Bache, at [redacted]@ucl.ac.uk or Professor Sutliffe, at [redacted]@ucl.ac.uk

| | |
|---|---|
| What is your relationship to your child with HIE? | ◯ Mother<br>◯ Father<br>◯ Other caregiver<br>◯ Other |
| If other, please specify | __________________________________ |
| Did your child receive cooling ("therapeutic hypothermia") in the UK? | ◯ Yes<br>◯ No<br>◯ Not sure |
| Was your baby transferred to a different hospital during or immediately after birth? | ◯ Yes<br>◯ No<br>◯ N/A |

| | |
|---|---|
| What year was your baby who experienced a HIE event born? | __________________________ |
| If you feel comfortable sharing, in which NICU(s) was your baby cared for in? | __________________________ |
| Which of these phrases best describes your child? | ◯ No developmental concerns or delays<br>◯ Developmental delays/issues<br>◯ Death from HIE<br>◯ Other<br>◯ Prefer not to say |
| Contact details | |
| What is your first name? | __________________________ |
| Are you willing to be contacted by email in relation to the study?<br>This will only be used for study related purposes e.g. for us to organise the interview and send you the Microsoft Teams joining link | ◯ Yes<br>◯ No |
| What is your email? | __________________________ |
| Please confirm your email | __________________________ |
| Which day(s) of the week would you prefer to do an interview online? Tick as many as apply | ☐ Monday<br>☐ Tuesday<br>☐ Wednesday<br>☐ Thursday<br>☐ Friday<br>☐ Saturday |
| What time of day would work best for an interview? Tick all that apply | ☐ Morning<br>☐ Afternoon<br>☐ Evening |
| What is your age? | ◯ 18 - 24<br>◯ 25 - 34<br>◯ 35 - 44<br>◯ over 44<br>◯ prefer not to say |
| What is your highest completed level of education? | ◯ secondary school<br>◯ college or A-levels<br>◯ university education<br>◯ other<br>◯ prefer not to say |
| How many children have you had? | ◯ 1<br>◯ 2<br>◯ 3 or more<br>◯ prefer not to say |

| | |
|---|---|
| Please choose your ethnicity | ◯ White British<br>◯ White - Irish<br>◯ White - Any other White background<br>◯ Mixed - White and Black Caribbean<br>◯ Mixed - White and Black African<br>◯ Mixed - White and Asian<br>◯ Mixed - Any other mixed background<br>◯ Asian or Asian British - Indian<br>◯ Asian or Asian British - Pakistani<br>◯ Asian or Asian British - Bangladeshi<br>◯ Asian or Asian British - Any other Asian background<br>◯ Black or Black British - Caribbean<br>◯ Black or Black British - African<br>◯ Black or Black British - Any other Black background<br>◯ Other Ethnic Groups - Chinese<br>◯ Other Ethnic Groups - Any other ethnic group<br>◯ Not known<br>◯ Prefer not to say |
| Do you have any questions about the interview process or study? We will respond by email if you have given permission | ________________________________ |

Please note by submitting this form you consent for us to store this data for the duration of the study. You may contact us to withdraw the data at any stage. We will not share your data with other third parties.

Notice:
The controller for this project is University College London (UCL). The UCL Data Protection Officer provides oversight of UCL activities involving the processing of personal data and can be contacted at. This 'local' privacy notice sets out the information that applies to this study. Further information on how UCL uses participant information can be found in our 'general' privacy notice, click here.
The categories of personal data used will be as follows:
Name, age, child's birth year, ethnicity, location
This will be used to ensure we listen to variety of families with different experiences, although you may choose not to share this information if you prefer, and you can still take part. The lawful basis that would be used to process your personal data will be performance of a task in the public interest. The lawful basis used to process special category personal data will be for scientific and historical research or statistical purposes. Your personal data will be processed so long as it is required for the research project. If we are able to anonymise or pseudonymise the personal data you provide we will undertake this and will endeavour to minimise the processing of personal data wherever possible. If you are concerned about how your personal data is being processed, or if you would like to contact us about your rights, please contact UCL in the first instance at.

[first_name], thank you so much for completing this. We really value your time and interest in helping this project.

3. Supplement 3 - Interview Guide

### Interview Guide

- Welcome participant and introduce self
- Confirm name of participant
- Ask if any questions
- Confirm that participant has had the chance to read and understand the information sheet and consent form
- Confirm consent form signed
- Explain the format of the interview and that participant is free to stop the interview at any stage or have a break. Explain that participants should only share information which they are comfortable to
- Check privacy and comfort
- Highlight support sheet with contacts and ability to contact researcher
- Request if happy to be recorded, explain about data withdrawal
- Begin recording if possible

***Appendix Figure 3-1. The interview guide questions.***

| **Question** | **Category probe** | **Detailed probe** |
|---|---|---|
| 1) *How has having a baby with HIE affected you and your family's lives? | | |
| | Emotional health | Worry, blame, guilt, embarrassment |
| | Mental health | PTSD |
| | | Depression |
| | | Anxiety |
| | Physical symptoms | Tiredness, sleep |
| | Social functioning | Energy<br>Childcare<br>Isolation |
| | Family relationships | With partner |
| | | With child's siblings |
| | | With grandparents |
| | Financial | Time off for appointments |
| | | Ability to work |

| | | |
|---|---|---|
| 2) *What have been the biggest struggles for your child<br>3) And for your family<br>4) and for yourself? | | |
| | Relational | Bonding<br>Conflict<br>Diversion of attention |
| | Clinical | Monitoring outcomes<br>Role and responsibility as a parent |
| | Emotional | Processing trauma + grief<br>Alienation/isolation |
| | Practical | Care<br>Travel |
| 5) What or who has been the greatest help or support? | | |
| | Friends and family | |
| | Support groups | |
| | Faith & beliefs | |
| 6) How have healthcare staff been helpful or not helpful? | | |
| 7) *What was the key information you were told on the neonatal unit about HIE? | | |
| | about what HIE is? | |
| | about the NICU & monitoring? | |
| | about cooling? | |
| | about MRI scans? | |

| | | |
|---|---|---|
| | about follow up and the future? | Feelings around preparedness for discharge<br>Role as parent |
| 8) How did you find these conversations? | | |
| | Communication of uncertainty? | |
| | And in the long run? | |
| | Would you change anything about these conversations? | |
| 9) *What would you change about the way you or your child were cared for? | | |
| | NICU environment | |
| | Communication | |
| | Staff interactions | |
| | Support after discharge | |

*Denotes key questions to be prioritised if time is short

- End recording
- Thank participant for taking part
- Debrief participant:
    - Ask if any questions
    - Check how feeling
    - Explain that participants can contact the research team and request for their data to be removed in the following week
    - Signpost participant to support sheet and organisations that can provide support
    - Inform participant that if they experience increased distress, they may contact one of the organisations listed in the where to get support sheet, additionally they are encouraged to contact the research team by email
    - According to the protocol of distress, the researcher may ask if the participant would like them to contact an organisation or personal contact for the participant, with their consent
    - Additionally, any follow up by the research team, as suggested by the Protocol of Distress, will be offered and confirmed

MRI – magnetic resonance imaging

4. Supplement 4 - Study Information

UCL
GREAT ORMOND STREET
INSTITUTE OF CHILD HEALTH

CHERUB

### The Impact of HIE on Families

#### What are we trying to do?

We aim to listen to families who have had a child with HIE in the UK to hear their experiences. We are interested in:

- what care was like on the neonatal unit
- support you received after discharge
- how the diagnosis affected you & your whole family.

We want to improve support and care for families affected by HIE in the UK.
This study is part of a Master of Research degree at UCL GOS Institute of Child Health (undertaken by Alexandra Bache, fifth year medical student). The supervisory team is based in the department of Population, Policy and Practice (Dr Rees, Professor Sutcliffe & Dr Lewis).

#### How will we do this?

We would like to speak to you and listen to your story ("interview" you). This will be online, on Microsoft Teams, at a time that works for you, and will last around one hour. You will be given a £20 voucher as a thank you for taking part. A trusted individual may join you for the individual interview, but we are particularly keen to hear your views.

#### Can I choose to take part?

Taking part is completely your choice. You can take part if you are:

- An adult aged 18 years or above
- And the **parent or caregiver of a baby who was diagnosed with and treated for hypoxic ischemic encephalopathy (HIE) with cooling in the UK**

You are not able to take part if you cannot understand English to a sufficient level to complete the study (as unfortunately translators are not available).

Please note, to ensure we listen to a variety of families, it may not be possible to interview all individuals who are interested. You are free to withdraw your participation from the study at any stage without giving a reason and you will be able to keep the £20 voucher. You may withdraw the data related to your interview within one week after the interview.

#### What will happen to my data?

Audio and videos of the interview will be recorded so that a transcript (written text of what you say) can be created. This will be "pseudo-anonymised". This means the data will be processed so that it will no longer be attributable to you unless additional information is available. This additional information (the recording) will only be available to the research team.

You may choose to turn off your video during the interview. The audio and video files will be safely stored and not available to anyone outside the research team and will be deleted at the end of the study (before the end of 2025). Direct quotes of what you say may be published in journals or used in conference presentations and lectures. However, these statements will be anonymised so that you cannot be identified (e.g. name and location changed). Anonymised transcripts may be stored for up to 5 years.

**Notice:** The sponsor for this project is University College London (UCL). The UCL Data Protection Officer provides oversight of UCL activities involving the processing of personal data and can be contacted at**.** This 'local' privacy notice sets out the information that applies to this study. Further information on how UCL uses participant information can be found in our 'general' privacy notice, click here. The categories of personal data used will be as follows: Name, Region, child's birth year, Ethnic origin

This will be used to ensure we listen to variety of families with different experiences, although you may choose not to share this information if you prefer, and you can still take part. The lawful basis that would be used to process your *personal data* will be performance of a task in the public interest. The lawful basis used to process *special category personal data* will be for scientific and historical research or statistical purposes. *Your personal data will be processed so long as it is required for the research project*. If we are able to anonymise or pseudonymise the personal data you provide we will undertake this and will endeavour to minimise the processing of personal data wherever possible. If you are concerned about how your personal data is being processed, or if you would like to contact us about your rights, please contact UCL in the first instance at.

#### What will happen to the interview findings?

Findings will be shared with the medical and scientific community through being published in a scientific journal (you will not be identifiable). To obtain a copy of the publication please contact [redacted]@ucl.ac.uk. The results will also be presented in other settings such as part of a Master of Research thesis and at conferences.

#### What are the risks & any support?

Taking part may carry some discomforts and risks, particularly:

- Uncomfortable & potentially distressing feelings and emotions
- Revisiting traumatic experiences

You will be provided with an information sheet detailing how to get help from relevant organisations. The research is in consultation with a child Bereavement Specialist. If at any stage you wish to stop the interview that decision will be respected and you will be signposted to further support, as outlined in the attached flowchart at the end.

Please note that confidentiality will be maintained as far as it is possible, unless during our conversation we hear anything which makes us worried that someone might be in danger of harm and not able to act for themselves. We might have to inform relevant agencies of this.

#### Complaints

If you would like to make a complaint or report an incident during the study please contact the Population Policy Practice Department at . If you feel your complaint has not been adequately handled, you can contact the Chair of the UCL Research Ethics Committee at

#### Who can I contact for more information?

Please contact Alexandra Bache ([redacted]@ucl.ac.uk) or Professor Sutcliffe by email, [redacted]@ucl.ac.uk). You will be given a copy of this information sheet and recommended to download a signed consent form to keep. **Thank you for reading this information sheet and for considering taking part in this research study.**

For your information, here is the flowchart we will be following to support you during the interview if you appear distressed:

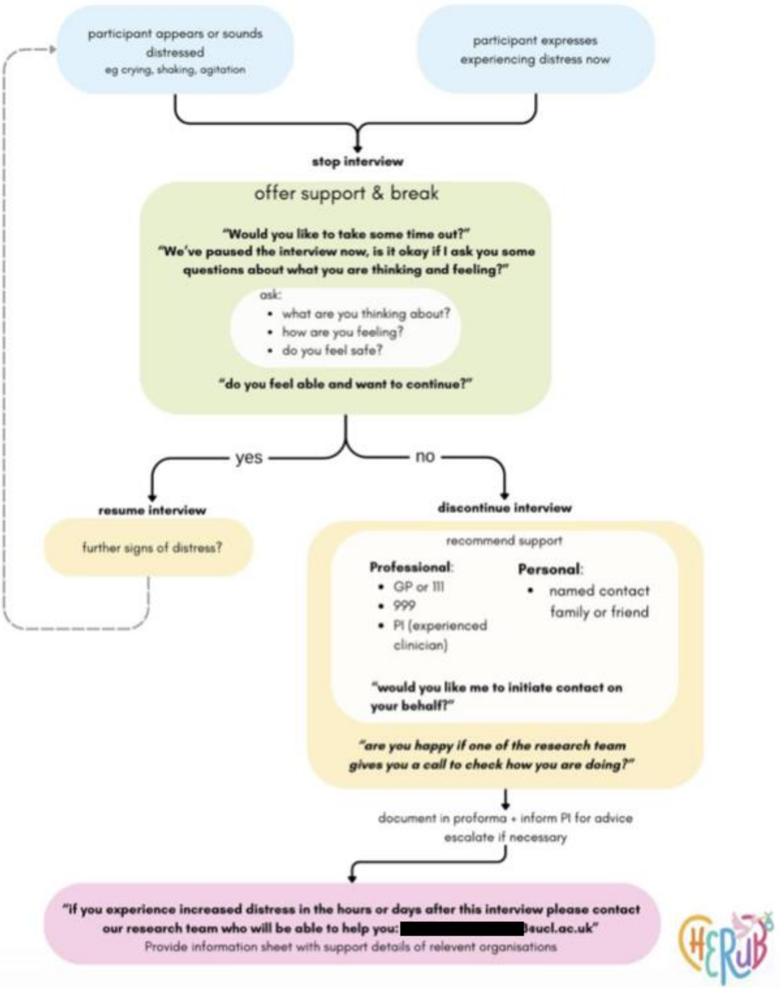

#### 5. Supplement 5 - E-consent

To see how this appeared to participants see Supplement 8-1B

### Interview Consent Form

Please complete this form after you have read the Information Sheet and/or listened to an explanation about the research.

Participant Information Sheet:

Participant Information Sheet.pdf

If you would like to ask any questions you can contact Alexandra Bache at [redacted]@ucl.ac.uk

Title of Study

Short title: The Impact of HIE on Families

Long title: The Impact of Neonatal Hypoxic Ischemic Encephalopathy on Families: A Mixed Methods Study

Department: Population Policy Practice, UCL Institute of Child Health

Name and Contact Details of the Researchers: Alexandra Bache; [redacted]@ucl.ac.uk Dr Philippa Rees; [redacted]@ucl.ac.uk [redacted]

Name and Contact Details of the Principal Researcher: Professor Alastair Sutcliffe; [redacted]@ucl.ac.uk

Name and Contact Details of the UCL Data Protection Officer: Alexandra Potts [redacted]@ucl.ac.uk

This study has been approved by the UCL Research Ethics Committee.

Project ID number: 26861/001

Thank you for considering taking part in this research. The person organising the research must explain the project to you before you agree to take part. If you have any questions arising from the Information Sheet or explanation already given to you, please ask the researcher before you decide whether to join in. You will be able to download a copy of this Consent Form to keep and refer to at any time.

I confirm that I understand that by clicking 'yes' for each box below I am consenting to this element of the study. I understand that clicking 'no' means that I DO NOT consent to that part of the study. I understand that by not giving consent for any one element that I may be deemed ineligible for the study.

For support:

Support available for you.pdf

| | | |
|---|---|---|
| 1) | I confirm that I have read and understood the Information Sheet for the above interview. I have had an opportunity to consider the information and what will be expected of me. I have also had the opportunity to ask questions which have been answered to my satisfaction and would like to take part in an individual interview. | ◯ Yes<br>◯ No |
| 2) | I understand that I will be able to withdraw my interview data within 1 week (7 days) after the interview | ◯ Yes<br>◯ No |

| | | |
|---|---|---|
| 3) | I consent to participate in the study. I understand that my personal information will be used for the purposes explained to me. I understand that according to data protection legislation, 'public task' will be the lawful basis for processing, and the lawful basis used to process special category personal data will be for scientific and historical research or statistical purposes. | ◯ Yes<br>◯ No |
| 4) | In the interviews all personal information will remain confidential.<br>All quotes will be presented anonymously. | ◯ Yes<br>◯ No |
| 5) | I understand that my information may be subject to review by responsible individuals from the University for monitoring and audit purposes. | ◯ Yes<br>◯ No |
| 6) | I understand that my participation is voluntary and that I am free to withdraw my participation at any time without giving a reason, without my legal rights being affected. | ◯ Yes<br>◯ No |
| 7) | I understand the potential risks of participating and the support that will be available to me should I become distressed during the research. | ◯ Yes<br>◯ No |
| 8) | No promise or guarantee of benefits, above and beyond a £20 thank you voucher, have been made to encourage me to participate | ◯ Yes<br>◯ No |
| 9) | I understand that the data will not be made available to any commercial organisations but is solely the responsibility of the researcher(s) undertaking this study. | ◯ Yes<br>◯ No |
| 10) | I understand that I will not benefit financially from this study or from any possible outcome it may result in in the future. | ◯ Yes<br>◯ No |
| 11) | I understand that I will still receive a £20 thank you voucher if I choose to withdraw. | ◯ Yes<br>◯ No |
| 12) | I agree that anonymised research data may be used by others for future research. [no one will be able to identify you when this data is shared] | ◯ Yes<br>◯ No |
| 13) | I understand that the information I have submitted will be published as a report | ◯ Yes<br>◯ No |
| 14) | I wish to receive a copy of the published report | ◯ Yes<br>◯ No |
| 15) | I consent to my interview being audio/video recorded and understand that the recordings will be:<br>-Stored securely on UCL servers<br>-Destroyed on completion of the study (before the end of 2025) | ◯ Yes<br>◯ No |

| | | |
|---|---|---|
| 16) | I hereby confirm that I understand the inclusion criteria as detailed in the Information Sheet and explained to me by the researcher. | ◯ Yes<br>◯ No |
| 17) | I hereby confirm that:<br>a)I understand the exclusion criteria as detailed in the Information Sheet and explained to me by the researcher; and<br>b)I do not fall under the exclusion criteria | ◯ Yes<br>◯ No |
| 18) | I am aware of who I should contact if I wish to lodge a complaint. | ◯ Yes<br>◯ No |
| 19) | I voluntarily agree to take part in an interview | ◯ Yes<br>◯ No |
| 20) | I understand the anonymous transcript may be archived for around 5 years | ◯ Yes<br>◯ No |

**Signature of participant**

| | | |
|---|---|---|
| 21) | First name | ________________________________ |
| 22) | Last name | ________________________________ |
| 23) | Signature of participant | ________________________________________ |
| 24) | Today's date | ________________________________ |

#### 6. Supplement 6 - Support Sheet

### SUPPORT AVAILABLE FOR YOU

We understand this can be a very difficult topic to talk about. It can bring up a whole range emotions. Here are some organisations you can contact before or following the interview if you experience distress or need support. You can also contact the research team.

##### emergency immediate help

eg suicidal thoughts, thoughts of harming someone else, you don't feel safe, get help now:

- **ring 999** or attend your local A&E
- contact a listening service:
  - call Samaritans on 116 123
  - text "SHOUT" to 85258 for text based support

##### emotional support

eg want peer support or HIE related resources

| Organisation name & website | What they offer |
|---|---|
| **Peeps HIE**<br>for those affected by HIE<br>https://www.peeps-hie.org | • HIE information<br>• peer support<br>• self-refer for more support<br>• counselling & trauma therapy |
| **Bliss**<br>for babies born premature or sick<br>https://www.bliss.org.uk | • email guidance<br>• Bliss facebook page<br>• information about neonatal care, going home & growing up<br>• volunteer video call if your baby is in hospital/recently discharged |
| **Together for Short Lives**<br>for seriously ill children & their families<br>https://www.togetherforshortlives.org.uk | • helpline - 08088088100<br>• family resources |
| **Birth Trauma Association**<br>for women and families who have experienced traumatic birth<br>https://www.birthtraumaassociation.org | • peer support eg Zoom drop ins, Facebook groups, helpline |

#### emotional support for grief

| Age of death | Organisation name & website | What they offer |
|---|---|---|
| Baby | **Petals**<br>the baby loss counselling charity<br>https://www.petalscharity.org | • one-to-one counselling in partnership with many hospital trusts |
| | **Sands**<br>supporting bereaved families<br>https://www.sands.org.uk | • helpline: 0808 164 3332<br>• support book<br>• online community<br>• support for groups such as men, siblings, long ago bereaved and Black and South Asian communities |
| Child | **Child Bereavement UK**<br>helps families to rebuild their lives when a child grieves or when a child dies<br>https://www.childbereavementuk.org | • helpline: 0800 02 888 40<br>• facilitated groups |
| | **The Compassionate Friends**<br>supporting bereaved parents and their families<br>https://www.tcf.org.uk | • online support<br>• retreats<br>• grief companions<br>• leaflets<br>• legal help |
| | **Care for the Family**<br>bereaved parent support<br>https://www.careforthefamily.org.uk | • befriending<br>• blog<br>• Facebook community<br>• support days |

#### 7. Supplement 7 - Distress Protocol

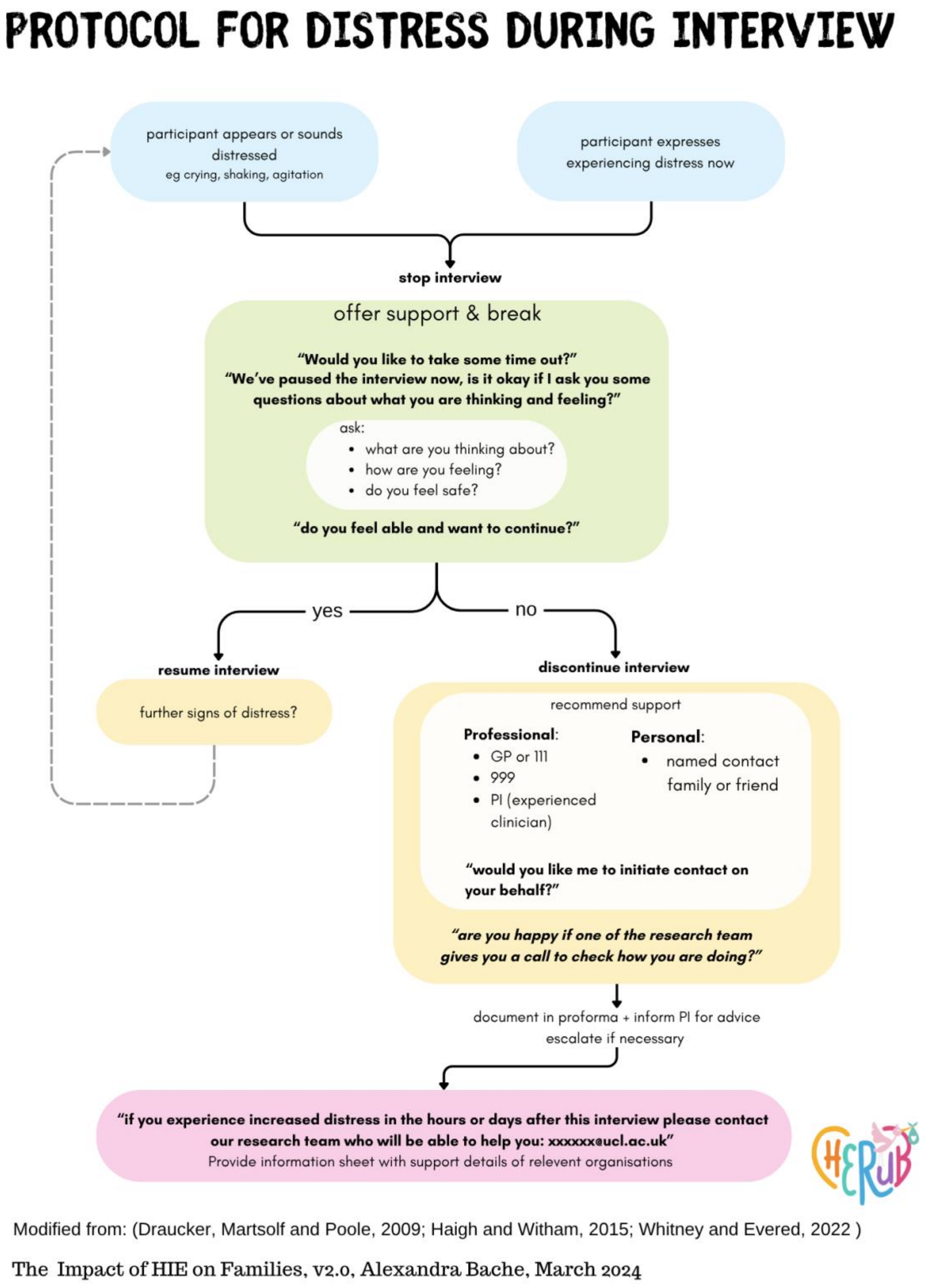

***Supplement Figure 7-1 Protocol for Distress.*** *See references on the next page.*

### REFERENCES

Draucker, C. B., Martsolf, D. S. and Poole, C. (2009) 'Developing Distress Protocols for Research on Sensitive Topics', Archives of Psychiatric Nursing, 23(5), pp. 343-350.

Haigh, C. and Witham, G. 2015. Distress Protocol for qualitative data collection. Manchester Metropolitan University

Whitney, C. and Evered, J. A. (2022) 'The Qualitative Research Distress Protocol: A Participant-Centered Tool for Navigating Distress During Data Collection', International Journal of Qualitative Methods, 21, pp. 16094069221110317.

#### 8. Supplement 8 - Study material Appearance

A

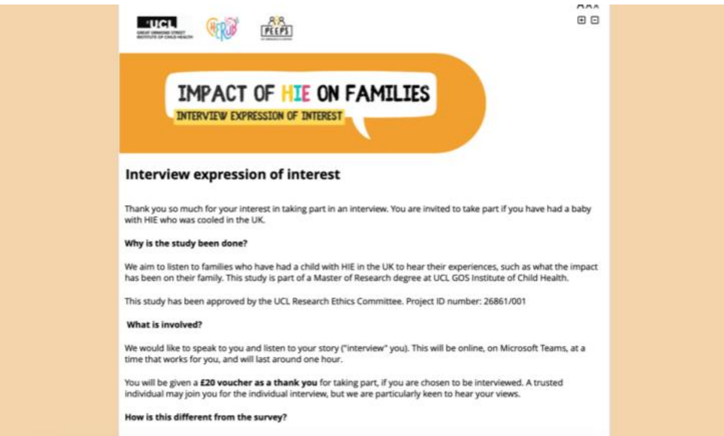

IMPACT OF HIE ON FAMILIES

INTERVIEW EXPRESSION OF INTEREST

**Interview expression of interest**

Thank you so much for your interest in taking part in an interview. You are invited to take part if you have had a baby with HIE who was cooled in the UK.

**Why is the study been done?**

We aim to listen to families who have had a child with HIE in the UK to hear their experiences, such as what the impact has been on their family. This study is part of a Master of Research degree at UCL GOS Institute of Child Health.

This study has been approved by the UCL Research Ethics Committee. Project ID number: 26861/001

**What is involved?**

We would like to speak to you and listen to your story ("interview" you). This will be online, on Microsoft Teams, at a time that works for you, and will last around one hour.

You will be given a **£20 voucher as a thank you** for taking part, if you are chosen to be interviewed. A trusted individual may join you for the individual interview, but we are particularly keen to hear your views.

**How is this different from the survey?**

B

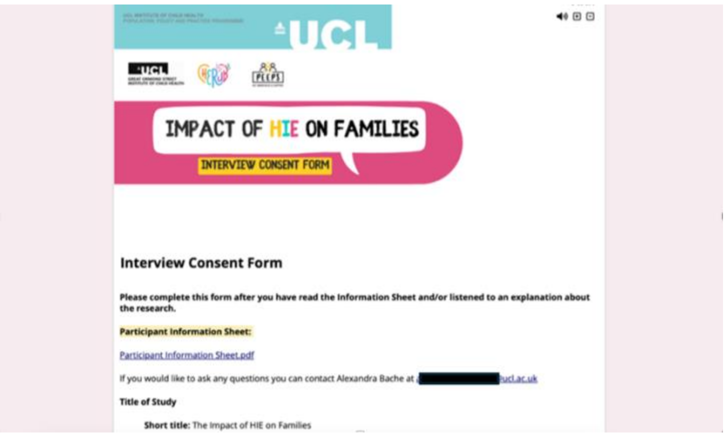

UCL

IMPACT OF HIE ON FAMILIES

INTERVIEW CONSENT FORM

**Interview Consent Form**

**Please complete this form after you have read the Information Sheet and/or listened to an explanation about the research.**

**Participant Information Sheet:**

Participant Information Sheet.pdf

If you would like to ask any questions you can contact Alexandra Bache at [redacted]ucl.ac.uk

**Title of Study**

**Short title:** The Impact of HIE on Families

***Supplement Figure 8-1. Screenshots to demonstrate how the questionnaires appeared to participants (A through B).*** *Interview expression of interest (A) and interview consent form (B) are shown.*
